# Calibration of Two Validated SARS-CoV-2 Pseudovirus Neutralization Assays for COVID-19 Vaccine Evaluation

**DOI:** 10.1101/2021.09.09.21263049

**Authors:** Yunda Huang, Oleg Borisov, Jia Jin Kee, Lindsay N. Carpp, Terri Wrin, Suqin Cai, Marcella Sarzotti-Kelsoe, Charlene McDanal, Amanda Eaton, Rolando Pajon, John Hural, Christine M. Posavad, Katherine Gill, Shelly Karuna, Lawrence Corey, M. Juliana McElrath, Peter B. Gilbert, Christos J. Petropoulos, David C. Montefiori

## Abstract

Vaccine-induced neutralizing antibodies (nAbs) are key biomarkers considered to be associated with vaccine efficacy. In United States Government-sponsored phase 3 efficacy trials of COVID-19 vaccines, nAbs are measured by two different validated pseudovirus-based SARS-CoV-2 neutralization assays, with each trial using one of the two assays. Here we describe and compare the nAb titers obtained in the two assays. We observe that one assay consistently yielded higher nAb titers than the other when both assays were performed on the World Health Organization’s anti-SARS-CoV-2 immunoglobulin International Standard, COVID-19 convalescent sera, and mRNA-1273 vaccinee sera. To overcome the challenge this difference in readout poses in comparing/combining data from the two assays, we evaluate three calibration approaches and show that readouts from the two assays can be calibrated to a common scale. These results may aid decision-making based on data from these assays for the evaluation and licensure of new or adapted COVID-19 vaccines.

## Introduction

Multiple vaccines have shown short-term efficacy against coronavirus disease-19 (COVID-19) in phase 3 randomized placebo-controlled trials^1-7^; one has been fully approved^8^ and three have been issued an Emergency Use Authorization^9^ by the US Food and Drug Administration (FDA), and six an Emergency Use Listing from the World Health Organization (WHO).^10^ To expediently meet the worldwide demand for sufficient doses (including booster doses) of safe and efficacious vaccines, and for the licensure of new or modified vaccines, the FDA and the WHO have recommended that immunobridging trials may be used to assess the effectiveness of booster vaccines, vaccines in new populations (e.g., pediatric) or against emerging SARS-CoV-2 variants,^11,12^ thus sidestepping the need for additional large-scale placebo-controlled randomized trials (which are becoming increasingly difficult to conduct, especially in high-income countries^13^) or active-control randomized trials.^14^ In such immunobridging trials, vaccine-induced immune responses of an established immune marker surrogate endpoint^15-17^ are typically assessed to ensure that levels deemed sufficient for clinical efficacy are achieved. Neutralizing antibodies (nAbs) are correlates of protection for many licensed vaccines,^17^ and an accumulating body of evidence^18-26^ supports their evaluation in immunobridging trials for COVID-19 vaccines.

In each of the five US Government (USG)-sponsored phase 3 trials, nAb responses are assessed by one of two different pseudovirus-based SARS-CoV-2 neutralization assays, one performed at Duke University and one at Monogram Biosciences. Each pseudovirus neutralization assay has been rigorously validated in accordance with the ICH/FDA guideline^27,28^ and these assays are planned for use in additional efficacy trials or immunobridging trials of COVID-19 vaccine candidates for regulatory approvals. If the Duke and Monogram assays do not render comparable readouts or are not calibrated (cross-validated) to a common scale, nAb response data from these assays may misguide regulatory decisions on these COVID-19 vaccine candidates.

To facilitate the harmonization of SARS-CoV-2 nAb data across multiple laboratories and assay types, the WHO developed an International Standard (WHO IS) for anti-SARS-CoV-2 immunoglobulin consisting of a pool of 11 COVID-19 convalescent plasma samples and provided as lyophilized material that, after reconstitution, was assigned a unitage of 1,000 international units per mL (IU/mL) for neutralization assays.^29^ Here we describe the Duke and Monogram nAb assays, and compare titer readouts between the two assays using a diverse set of samples that include the WHO IS, COVID-19 convalescent serum samples, and serum samples from Moderna mRNA-1273 vaccine recipients. Because the ultimate goal is to ensure comparability of nAb readouts in vaccine recipients, we also present and compare statistical approaches for calibrating readouts from the two assays to support future vaccine evaluation and development.

## Results

Table 1 summarizes and compares the Duke and Monogram assays. Similarities of both assays include utilizing lentiviral vector that 1) is pseudotyped with full-length G614 Spike protein (the SARS-CoV-2 surface protein responsible for virus entry via interaction with the human ACE2 receptor^30,31^) and 2) contains a firefly luciferase (Luc) reporter gene. Thus, in both assays a quantitative luminescence readout is directly proportional to the amount of pseudovirus entry into target cells. In both assays, neutralizing antibodies in the assayed sample inhibit pseudovirus entry and reduce luminescence signal in a dose-dependent manner, providing the basis for quantifying the neutralizing activity of the sample. Both assays are established to measure ID50 and ID80 neutralizing titers and are fully validated in terms of all required assay performance parameters^27^ for convalescent serum and partially validated in terms of specificity, linearity, precision and limits of quantitation of the assay for vaccine incurred samples, in order to extend validation for the purpose of testing clinical trial specimens. Differences in pseudovirus preparation, cell lines, plate layouts, and dilution schemes are also noted in Table 1. Among the notable differences is the way that Transmembrane Protein Serine 2 (TMPRSS2) is introduced into the assay. SARS-CoV-2 fuses with host cells by binding to the ACE2 receptor; the host cell TMPRSS2 protease cleaves the Spike protein at the S1/S2 sites to facilitate fusion.^32^ In the Duke assay, TMPRSS2 is expressed during pseudovirus production, whereas in the Monogram assay it is expressed on the surface of HEK 293T/ACE2 target cells. This difference can impact relative neutralization titers measured in these assays, with higher titers seen in HEK 293T/ACE2/TMPRSS2 cells (Montefiori, unpublished).

**Table 1.**
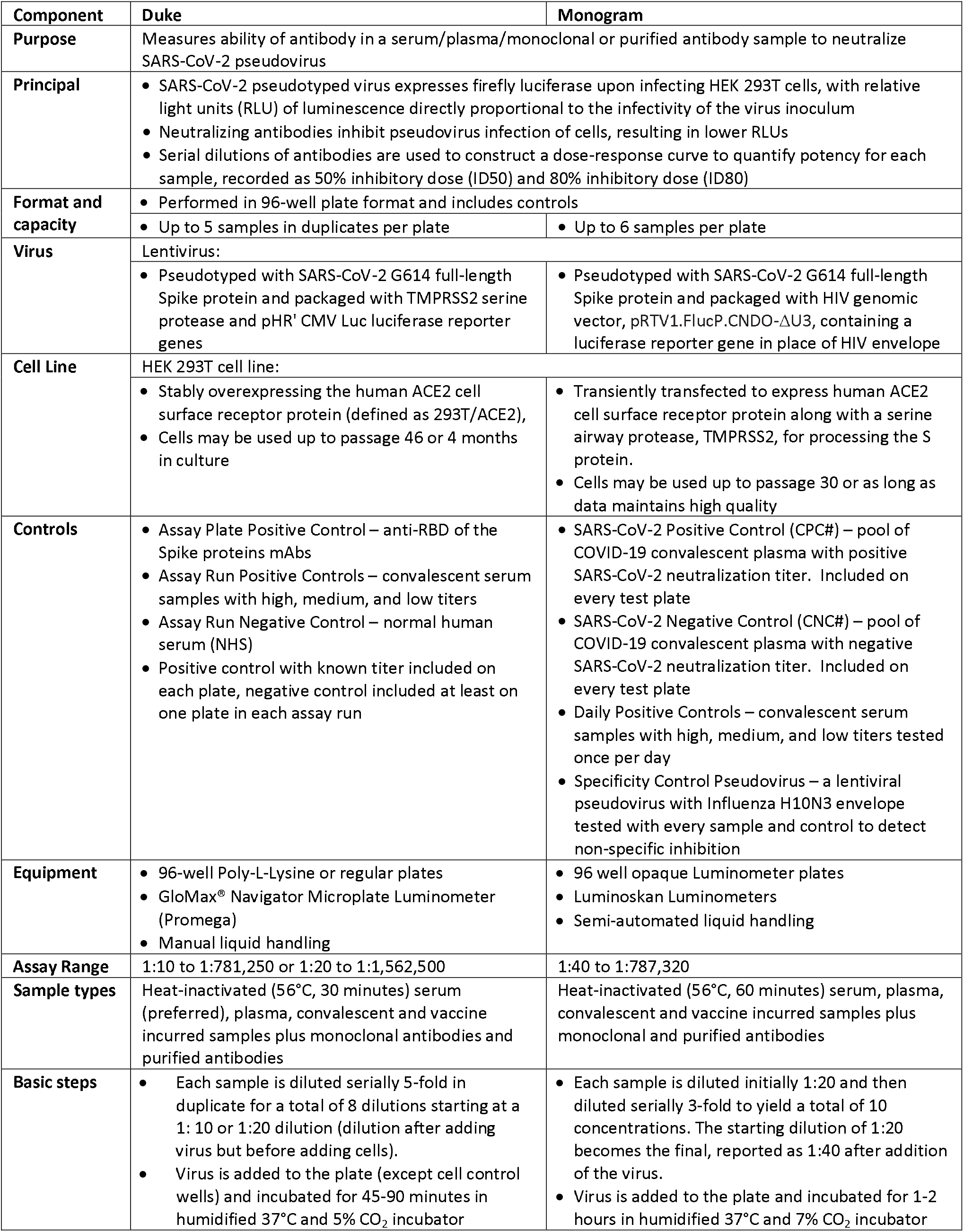
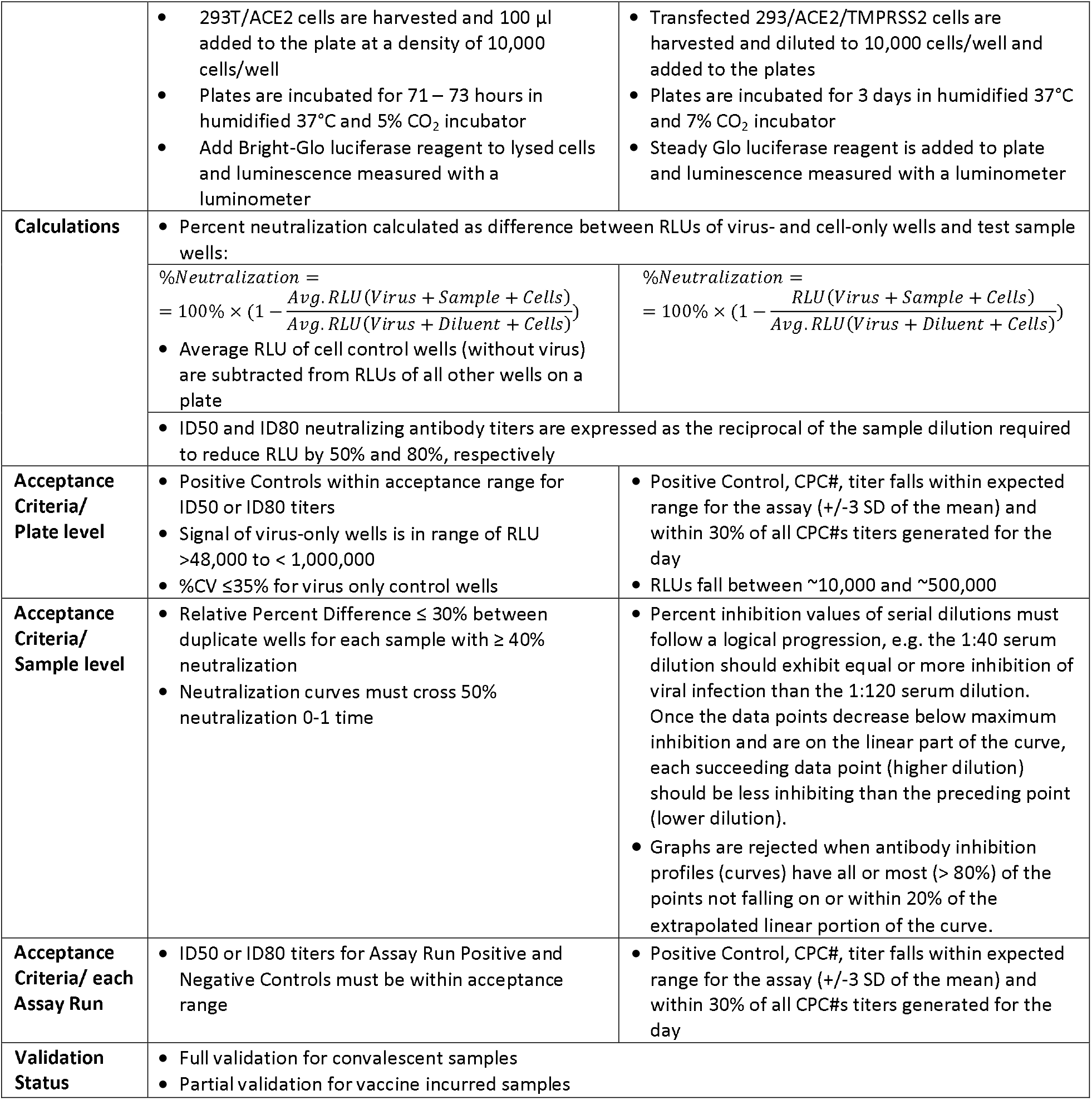
Overview comparison of the Duke vs. Monogram assays.

For assay comparison and establishment of the calibration algorithms, nAbs were measured in the first WHO IS for anti-SARS-CoV-2 immunoglobulin (20/136), as well as in 248 convalescent serum samples by both the Duke and Monogram labs following each assay format. For demonstration of the performance of the proposed calibration methods, nAbs were measured by both labs in 90 mRNA-1273 vaccine recipient serum samples [30 participants at three time points each: Day 1 (baseline), Day 29 (4 weeks post first vaccination), and Day 57 (4 weeks post second vaccination)].

Among the 90 samples from vaccine recipients, all Day 1 (pre-vaccination) samples had non-detectable nAbs by both labs, indicating a high level of specificity of the assays. Moderate nAb titers were detected after the first vaccination at Day 29 among almost all of the participants for ID50 (30 out of 30, 100% by Duke, 27 out of 30, 90% by Monogram) and the majority of participants for ID80 (80% by Duke, 70% by Monogram). nAb titers at Day 57 were detected among all participants who received the second dose (100% for ID50 and ID80 by both labs), and Day 57 titers were on average more than 20-fold higher than Day 29 nAb responses in both assays (Figure 1, Table 2). Specifically, geometric mean ID50 titers (standard deviation of log_e_-transformed titers, SD) were 65.7 (0.8) at Day 29 and 1463.5 (1.1) at Day 57 as measured by the Duke lab and 178.3 (1.0) at Day 29 and 4120.7 (1.0) at Day 57 as measured by the Monogram lab. Geometric mean ID80 titers were 16.2 (0.8) at Day 29, and 330.5 (0.9) at Day 57 as measured by the Duke lab; 60.9 (0.8) at Day 29 and 1522.5 (1.0) at Day 57 as measured by the Monogram lab. Moderate correlations were seen between the Day 29 and Day 57 nAb titers (Figure S1). Day 29 and Day 57 nAb titers were treated separately in the calibration process, in order to cover different dynamic ranges of possible assay readouts.

**Table 2:**
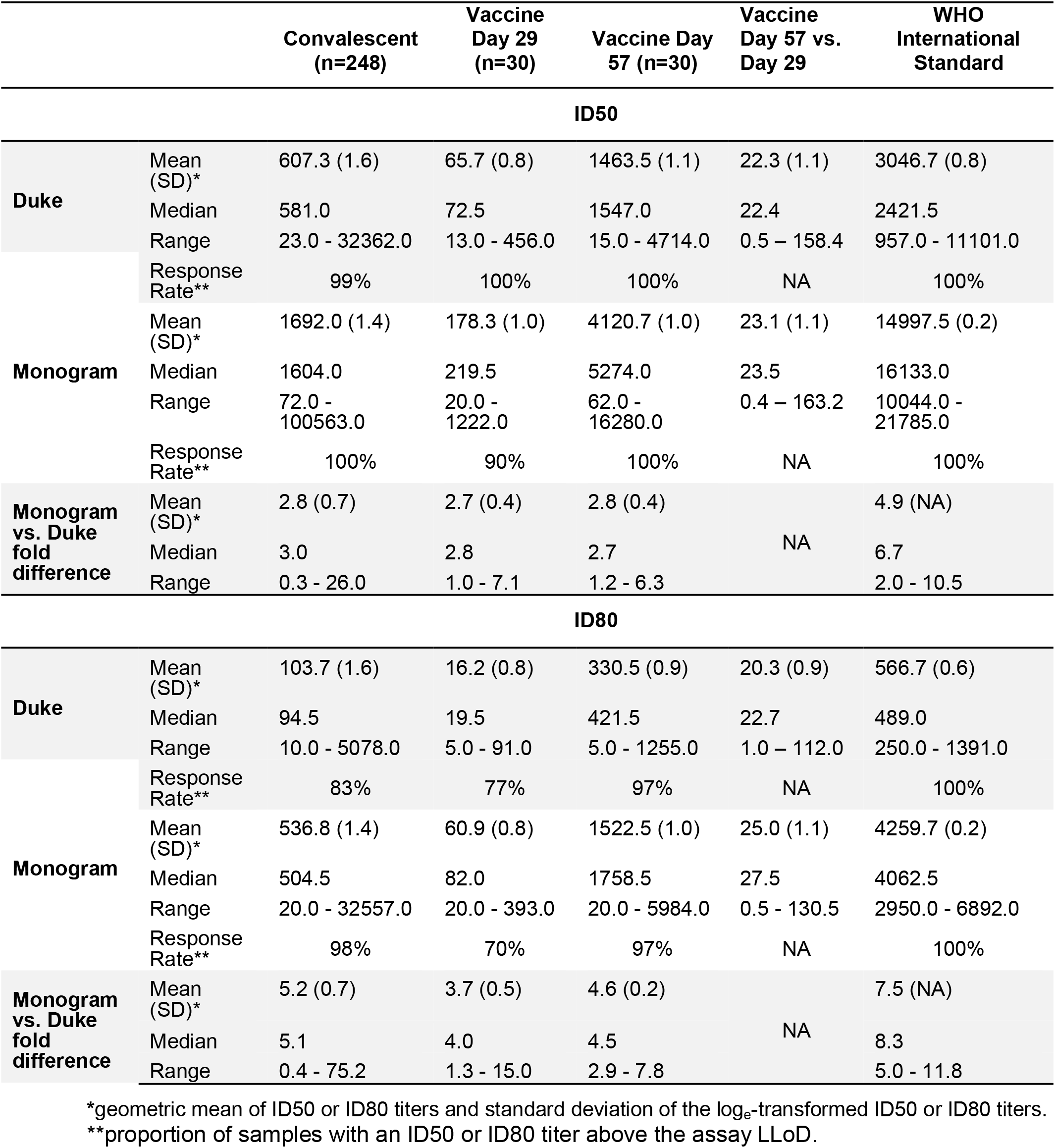
Summary of ID50 and ID80 titers from each lab and their fold-differences for convalescent patient sera, Day 29 vaccinee sera, Day 57 vaccinee sera, and the WHO International Standard sample.

**Figure 1:**
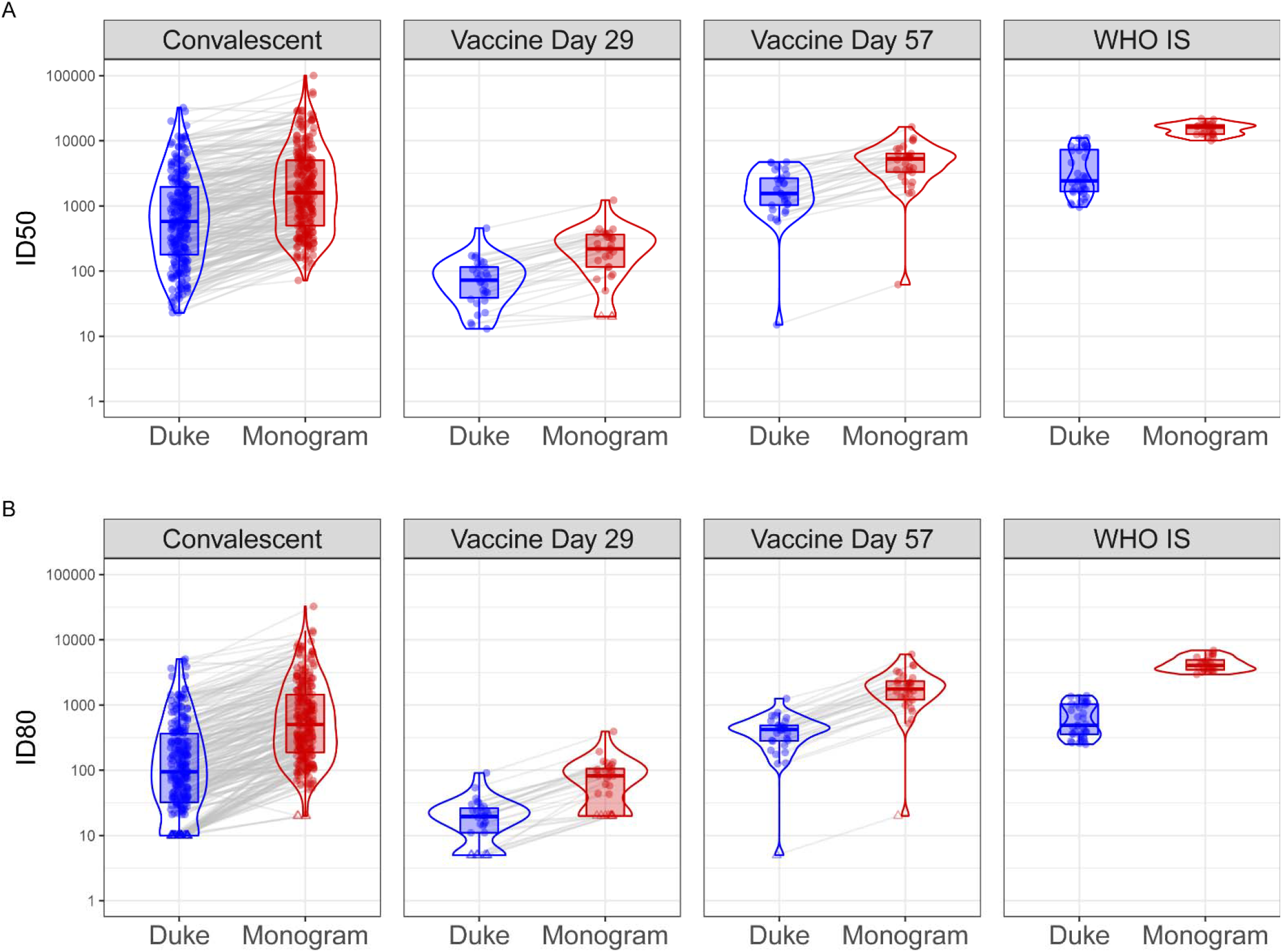
Distributions of ID50 (Panel A) and ID80 (Panel B) titers measured by the Duke and Monogram labs of convalescent patient samples, vaccine recipient samples, and the WHO International Standard sample. For the convalescent samples, the assay lower limit of detection (LLoD) is 1:20 for Duke and 1:40 for Monogram. For the vaccine samples collected at Day 29 and Day 57, 4 weeks post the first and the second Moderna doses, respectively, and the WHO IS sample, the assay LLoD is 1:10 for Duke and 1:40 for Monogram. Circle-shape points indicate titers > LLoD; triangle-shape points indicate titers ≤ LLoD.

Prior to calibration, we observed that ID50 and ID80 titer readouts from the Monogram lab were consistently higher than those from the Duke lab measured in convalescent sera, vaccine recipient sera, and the WHO IS sample (Figure 1, Table 2). Specifically, the fold difference of individual-level ID50 titers from Monogram vs. Duke across individuals had a geometric mean (SD) of 2.8 (0.7) in convalescent samples, 2.7 (0.4) and 2.8 (0.4) in Day 29 and Day 57 vaccine recipient samples, respectively, and the fold difference of the geometric (arithmetic) mean ID50 titers across replicates of the WHO IS sample was 4.9 (3.7); the fold difference of individual-level ID80 titers Monogram vs. Duke across individuals had a geometric mean (SD) of 5.2 (0.7) in convalescent samples, 3.7 (0.5) and 4.6 (0.2) in Day 29 and Day 57 vaccine recipient samples, respectively, and the fold difference of the geometric (arithmetic) mean ID80 titers across replicates of the WHO IS sample was 7.5 (6.6). These differences in readouts between the two assays are an example of the need for calibration of the responses to the same scale prior to cross-assay comparisons or cross-assay merging of results. The fact that fold differences between the two assays are largely consistent across convalescent samples and vaccine recipient samples suggests that the differences between the assays are not specific to naturally acquired or vaccine-elicited nAbs. In addition, the range of readouts in convalescent samples generally covers that in vaccine recipient samples; this supports the use of those convalescent samples to develop the algorithms for the calibration of titers in the vaccine recipient samples. On the other hand, the WHO IS, which is a single sample comprised of a pool of convalescent plasma samples, does not cover the entire range of titers and hence resulted in an expected narrower range of replicate values.

We evaluated three different approaches for the calibration of vaccine-induced neutralizing antibody response titers measured by the Duke and Monogram labs. Approach 1 uses tiers of the WHO IS sample to ‘standardize’ nAb titers measured by each lab to a common international unit (IU/ml),^33^ whereas Approaches 2 and 3, via two different statistical algorithms (details in Methods), use titers of an independent pool of clinical (e.g. convalescent) samples measured by both labs to calibrate titers measured by each lab to a common scale still in titer units.

For Approach 1, calibration factors were calculated using the arithmetic mean, geometric mean or median titers of the WHO IS sample measured by both labs (Table S1). Using the arithmetic mean-based calibration factor, calibrated ID50 (cID50) and ID80 (cID80) readouts of the Day 29 and Day 57 vaccine-induced responses from the two labs rendered the highest agreement, compared to the geometric- and median-based factors, with a concordance correlation coefficient (CCC) of 0.75 (95% CI: 0.60, 0.85) for cID50, and 0.72 (0.56, 0.82) for cID80 (Figure S2). Therefore, Approach 1 using the arithmetic mean-based calibration factor is used to compare with Approaches 2 and 3 (Figure 2).

**Figure 2:**
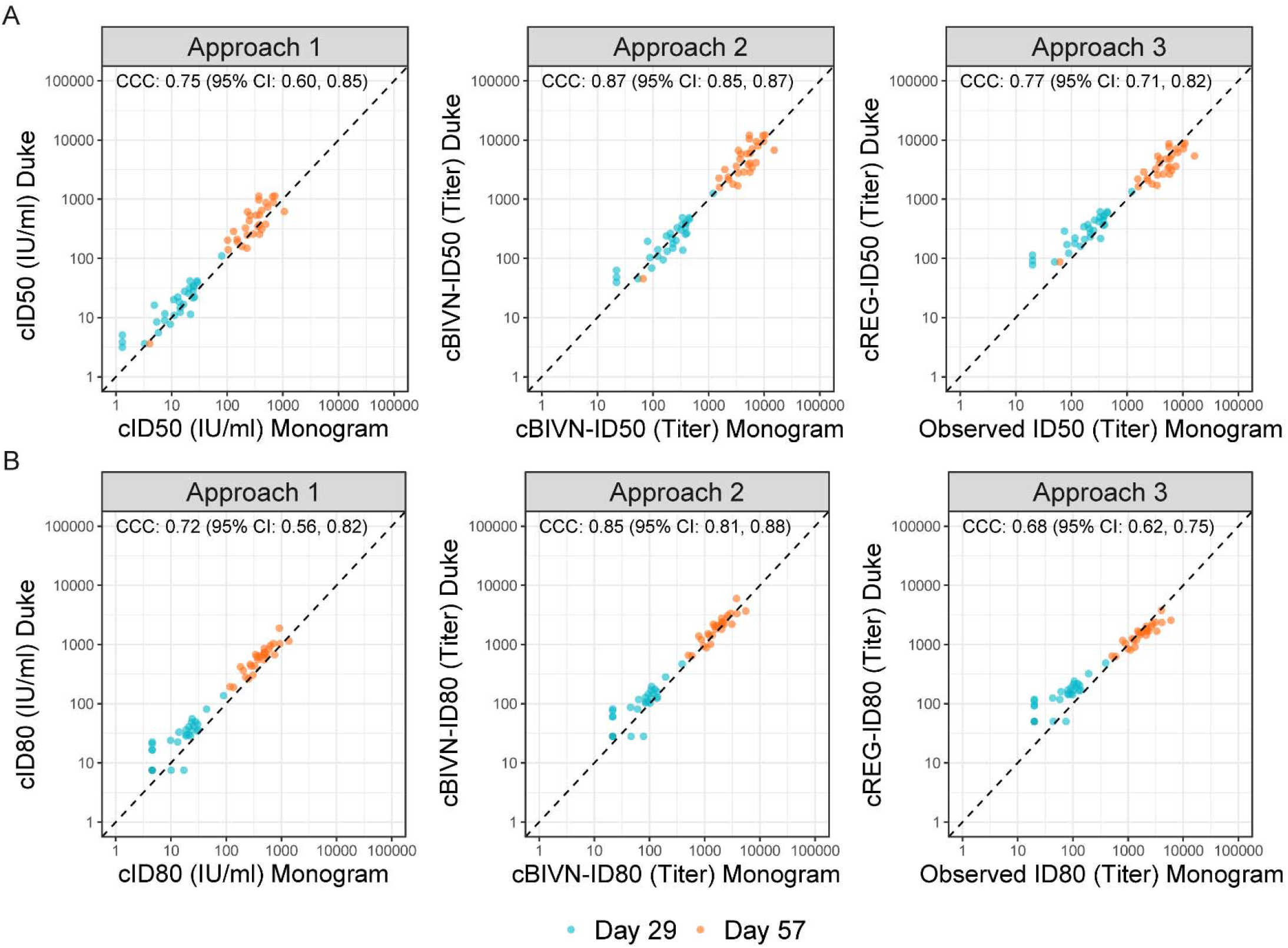
Scatterplots of calibrated neutralizing antibody readouts demonstrating performance of the three calibration approaches using vaccine recipient samples collected at Day 29 (turquoise) and Day 57 (orange), 4 weeks post the first and the second mRNA-1273 vaccine doses, respectively. The arithmetic mean-based calibration factor was used in Approach 1. The concordance correlation coefficient (CCC) and 95% confidence intervals indicate the level of agreement between the x- and y-axis values. • cID50, cID80: ID50, ID80 titers calibrated to the WHO International Standard, expressed in International Units per ml (IU/ml) (Approach 1). • cBIVN-ID50, cBIVN-ID80: ID50, ID80 titers from each lab calibrated to a common scale using an independent pool of clinical samples, assuming a bivariate normal distribution for the readouts from the two labs (Approach 2). • cREG-ID50, cREG-ID80: ID50, ID80 titers from the non-reference lab calibrated to the reference lab using an independent pool of clinical samples, based on regressing titers from the reference lab on the non-reference lab (Approach 3).

For Approach 2, readouts from the two labs were calibrated to the same scale (with the Monogram readouts set to the reference) via an algorithm developed using titers of the convalescent samples. This approach assumes a bivariate normal (BIVN) distribution of the two assay readouts and incorporates the reported measurement error of each assay. Calibrated vaccine-induced neutralizing antibody ID50 and ID80 titers (cBIVN-ID50 and cBIVN-ID80, respectively) using Approach 2 achieved the highest agreement among the three approaches with a CCC of 0.87 (0.85, 0.87) for cBIVN-ID50 titers, and 0.85 (0.81, 0.88) for cBIVN-ID80 titers.

Similar to Approach 2, readouts from the Duke lab in Approach 3 were calibrated to the scale of the Monogram lab via an algorithm also developed using titers of the convalescent samples. Unlike Approach 2, Approach 3 is based on regressing (REG) the log-transformed titers of the Monogram assay (reference lab) on the Duke assay without incorporating assay measurement errors (Figure S3). Calibrated ID50 and ID80 titers (cREG-ID50 and cREG-ID80) from the Duke lab and observed readouts from the Monogram lab also showed reasonable agreement with a CCC of 0.77 (0.71, 0.82) for ID50 titers, and 0.68 (0.61, 0.75) for ID80 titers, comparable to the performance of Approach 1.

## Discussion

Neutralizing antibody data obtained from multiple trials of different vaccines can support the establishment of nAbs as a correlate of protection, and if demonstrated, may facilitate approval of future vaccines through immunobridging. It is not often possible for samples from different vaccine trials to be evaluated in a single assay and laboratory. In the case of SARS-CoV-2, nAb data were generated at one of two independent laboratories for each of the five USG-sponsored vaccine efficacy trials; this approach is planned for future trials, as well. Because assay readouts in vaccine recipient sera from these two laboratories are on average 3-5 fold different, our study addresses this imperative need via calibrating data from the two assays to the same scale.

Our first main conclusion is that the WHO IS sample (Approach 1) demonstrated satisfactory performance comparable to that of the panels of convalescent sera in calibrating COVID-19 vaccine-induced nAb titers measured by these laboratories. This finding has important implications for future development and evaluation of COVID-19 vaccines because the WHO IS is more widely available and hence provides a straightforward way to compare and combine data across different platforms, and/or different developers and laboratories. Another main conclusion is that any of the three different calibration approaches can be used to bring readouts from different assays to the same scale for further analyses across studies, but that, due to the distinct advantages and disadvantages of each approach (summarized in Table S2), the specific approach that will be best suited for a given analysis depends on the availability of data to establish the calibration algorithms. For our motivating application – cross-protocol immunogenicity and correlates analyses of the 5 USG COVID-19 trials, where all nAb data are obtained by either the Duke or the Monogram assay – Approach 2 is recommended. However, for analyses including nAb data not measured by either the Duke or Monogram assay, then Approach 1 (using International Units) is likely the most feasible option, because not all labs have access to the convalescent samples that are necessary for Approach 2. We discuss other implementation considerations for the three Approaches below.

Among the three approaches, Approach 1 is the easiest to implement and likely requires the fewest resources as long as data on the WHO IS sample are available from the lab from a sufficient number of vials (e.g., > 20-30) and replicates. One requirement is to choose an appropriate summary statistic to calculate the calibration factor. In the dataset presented in this study, the calibration factor based on the arithmetic mean of ID50 or ID80 titers rendered the best performance. This is largely due to the fact that the corresponding WHO standard-based calibration factor for each lab translates to conversion factors, i.e., the ratio of readouts in international units of 3.7 for ID50 and 6.6 for ID80 titers between Monogram vs. Duke labs, that are in line with the fold-differences observed in the convalescent patient samples and in the vaccine recipient samples. This explains why this simple multiplicative-factor approach rendered reasonable calibration performance. Meanwhile it also highlights that external neutralizing antibody titers from convalescent or vaccine sera generated by both labs on a common set of samples can inform decisions on which summary statistic to use in Approach 1 by evaluating the conversation factor against the additional data sources. Approach 1 is currently being used in the analyses of nAb data generated from two of the USG trials, one set of data from the Duke lab and the other by the Monogram lab. These analyses provide an important validation that assay-specific differences are minimized in the resulting calibrated data based on Approach 1 across the two USG trials, so that potential differences or similarity between the vaccine candidates in terms of their immune responses are not masked by differences between the two assays.

For both Approaches 2 and 3, a common pool of convalescent or vaccine samples will need to be assayed by both labs in order to establish the calibration algorithm, where Approach 2 requires readouts from both labs to be calibrated to a common scale, and Approach 3 calibrates the readouts from one lab to the other reference lab. We find that Approach 2 rendered the best calibration performance, followed by comparable performance between Approaches 1 and 3 for both ID50 and ID80 titers. These results suggest that precision in calibration can be improved by accounting for assay measurement error and the differences between the two assays across a range of response magnitudes (i.e., as done in Approach 2). In our case, measurement errors were characterized in assay validation experiments using both convalescent samples and vaccine samples by both labs, and assay differences in magnitude were characterized using convalescent samples with dynamic ranges of titers that were comparable to the ranges of titers of vaccine trial samples requiring calibration. Therefore, Approach 2 is recommended when paired-sample data are available from the two assays/labs (as is the case for the analyses of assay data from the 5 USG COVID-19 vaccine trials, where both the Duke and Monogram labs assayed a common set of convalescent samples for the calibration of their readouts in vaccine samples).

There are several limitations of this study. First, for evaluating the performance of the calibration approaches, we only considered a limited number of samples from trials of the mRNA-1273 vaccine. This potentially limits our ability to generalize findings from the calibration study to other COVID-19 vaccine platforms. We plan to assay a subset of samples from phase 3 trials of other COVID-19 vaccines to further validate the calibration approaches when those data become available. Second, to develop the calibration algorithms, both labs must have their assays performed on a common pool of independent samples that are expected to cover a similar dynamic range as the readouts requiring calibration. Access to such a common pool may be challenging for many labs. In that case, Approach 1 might be the most feasible option as long as the WHO IS sample is available in sufficient quantity. Third, for all three approaches, we focused the calibration only for titers above the lower limit of detection (LLoD) of the reference assays. Methods that account for calibration of responses below the LLoD, especially in the context of two assays having different LLoDs, are currently under development. Fourth, this study only concerns data from two specific labs measuring nAbs. Further validation is needed to investigate the performance of the three approaches applied to the calibration of other assays and/or immune responses. Although the described calibration approaches are only applied to the calibration of assays measuring anti-SARS-CoV-2 nAb responses induced by candidate vaccines, a more general use is amenable when assay data from multiple sources are of interest for comparison or combination. Fifth, the two nAb assays used in this study were both pseudovirus-based using lentiviral packaging. Future studies could also compare nAb responses obtained by the pseudovirus assay to those obtained by other SARS-CoV-2 neutralization platforms, e.g., pseudovirus-based neutralization using other viral vectors, or neutralization of live SARS-CoV-2. Lastly, this study focused on a single variant of SARS-CoV-2 (G614) that dominated the pandemic during the earliest phase 3 trials (e.g., Pfizer and Moderna trials), but has since been replaced by other variants, most recently the Delta variant.^34^ Although the G614 form of Spike remains an important comparator, additional efforts are needed to establish new standards for current and future SARS-CoV-2 variants of concern.

Our study has a number of strengths. First, three distinct statistical approaches for calibration, each with different data requirements and/or data assumptions, are discussed to provide researchers versatile tools to help guide decisions for which approach may be optimal for calibrating a given data set. Second, each assay was formally validated for the assessment of COVID-19 vaccine-induced or naturally acquired neutralizing antibody immune responses, according to current regulatory guideline and standards. Third, to our knowledge, this is the first study that uses convalescent sera and the WHO IS sample to establish calibration algorithms and evaluate the performance of different calibration approaches for samples from COVID-19 vaccine trials. When more data become available, each approach could be readily updated to improve calibration accuracy and precision. Lastly, although the described calibration approaches were only illustrated for the analysis of data from two labs, all three approaches can be easily generalized to the calibration of data from more than two assays or labs.

In summary, correlates and immuno-bridging studies that could accelerate the development and evaluation of new and modified vaccines are at the heart of COVID-19 prevention efforts. We presented calibration approaches to support valid comparisons and combination of key immune response data from these studies to minimize the influence of assay-specific differences and optimize the characterization of future vaccine candidates.

## Online Methods

### Pseudovirus neutralization assays

Table 1 provides a comparison of the assays’ components and major steps. Further details are provided for each assay below.

#### Duke

Neutralization was measured in a formally validated assay that utilized lentiviral particles pseudotyped with full-length SARS-CoV-2 Spike protein and containing a firefly luciferase (Luc) reporter gene for quantitative measurements of infection by relative luminescence units (RLU). The assay was performed in 293T/ACE2.MF provided by Drs. Michael Farzan and Huihui Mu. Pseudoviruses were prepared and titrated for infectivity essentially as described previously.^35^ Briefly, an expression plasmid encoding codon-optimized full-length spike of the Wuhan-1 strain (VRC7480) was provided by Drs. Barney Graham and Kizzmekia Corbett at the Vaccine Research Center, National Institutes of Health (USA). Mutations, including the D614G mutation, were introduced into VRC7480 by site-directed mutagenesis^35^ using the QuikChange Lightning Site-Directed Mutagenesis Kit from Agilent Technologies (Catalog # 210518). All mutations were confirmed by full-length spike gene sequencing by Sanger Sequencing, using Sequencher and SnapGene for sequence analyses. Pseudovirions were produced in HEK293T/17 cells (ATCC cat. no. CRL-11268) by transfection using Fugene 6 (Promega Cat#E2692) and a combination of spike plasmid, lentiviral backbone plasmid (pCMV ΔR8.2) and firefly Luc reporter gene plasmid (pHR’ CMV Luc)^36^ in a 1:17:17 ratio in Opti-MEM (Life Technologies). Transfection mixtures were added to pre-seeded HEK 293T/17 cells in T-75 flasks containing 12 ml of growth medium and incubated for 16-20 h at 37°C. Medium was removed and 15 ml of fresh growth medium added. Pseudovirus-containing culture medium was collected after an additional 2 days of incubation and clarified of cells by low-speed centrifugation and 0.45 µm micron filtration. TCID50 assays were performed as described previously.^35^ For measurements of neutralization, a pre-titrated dose of virus was incubated with 8 serial 5-fold dilutions of serum samples (1:10 or 1:20 starting dilution) in duplicate in a total volume of 150 µl for 1 h at 37°C in 96-well flat-bottom poly-L-lysine-coated culture plates. 293T/ACE2-MF cells were detached from T75 culture flasks using TrypLE Select Enzyme solution, suspended in growth medium (100,000 cells/ml) and immediately added to all wells (10,000 cells in 100 µL of growth medium per well). One set of 8 wells received cells + virus (virus control) and another set of 8 wells received cells only (background control). After 66-72 h of incubation, medium was removed by gentle aspiration and 30 µl of Promega 1X lysis buffer was added to all wells. After a 10-minute incubation at room temperature, 100 µl of Bright-Glo luciferase reagent was added to all wells. After 1-2 minutes, 110 µl of the cell lysate was transferred to a black/white plate. Luminescence was measured using a GloMax Navigator luminometer (Promega). Serum samples were heat-inactivated for 30 minutes at 56°C prior to assay. Neutralization titers are the inhibitory dilution (ID) of serum samples at which RLUs were reduced by either 50% (ID50) or 80% (ID80) compared to virus control wells after subtraction of background RLUs.

#### Monogram

Neutralizing antibody activity was measured in a formally validated assay that utilized lentiviral particles pseudotyped with full-length SARS-CoV-2 Spike protein and containing a firefly luciferase (Luc) reporter gene for quantitative measurements of infection by relative luminescence units (RLU). The backbone vector used in pseudovirus creation, F-lucP.CNDOΔU3, encodes the HIV genome with firefly luciferase replacing the HIV *env* gene. A codon-optimized version of the full-length spike gene of the Wuhan-1 SARS-CoV-2 strain (MN908947.3) (GenScript) was cloned into the Monogram proprietary *env* expression vector, pCXAS-PXMX, for use in the assay. The D614G spike mutation was introduced into the original Wuhan sequence by site-directed mutagenesis. Sequences of the spike gene and expression vector were confirmed by full-length sequencing using Illumina MiSeq NGS.

Pseudovirus stock was produced in HEK 293 cells via a calcium phosphate transfection using a combination of spike plasmid (pCXAS-SARS-CoV-2-D614G) and lentiviral backbone plasmid (F-lucP.CNDOΔU3). Transfected 10 cm^2^ plates were re-fed the next day and harvested on Day 2 post transfection. The pseudovirus stock (supernatant) was collected, filtered and frozen at ≤70°C in single-use aliquots. Pseudovirus infectivity was screened at multiple dilutions using HEK293 cells transiently transfected with ACE2 and TMPRSS2 expression vectors. RLUs were adjusted to ∼50,000 for use in the neutralization assay. Neutralization was performed in white 96-well plates by incubating pseudovirus with 10 serial 3-fold dilutions of serum samples for one hour at 37°C. Serum samples were heat-inactivated for 60 minutes at 56°C prior to assay.

The dilution series was based on a 1:20 starting dilution which was reported as 1:40 after addition of virus. HEK293 target cells, which had been transfected the previous day with ACE2 and TMPRSS2 expression plasmids, were detached from 10 cm^2^ plates using trypsin/EDTA and re-suspended in culture medium to a final concentration that accommodated the addition of 10,000 cells per well. Cell suspension was added to the serum-virus mixtures and assay plates were incubated at 37°C in 7% CO_2_ for 3 days. On the day of assay read, Steady Glo (Promega) was added to each well. Reactions were incubated briefly and luciferase signal (RLU) was measured using a luminometer. Neutralization titers represent the inhibitory dilution (ID) of serum samples at which RLUs were reduced by either 50% (ID50) or 80% (ID80) compared to virus control wells (no serum wells).

The Monogram assay employs a specificity control which is created using the same HIV backbone/Luc sequence used in the SARS-CoV-2 pseudovirus. The envelope is 1949 Influenza A H10N3. It is unlikely for human sera to have antibodies against this rare avian influenza virus. The specificity control is designed to detect non-antibody factors (e.g., ART therapy) that could inhibit SARS-CoV-2 pseudovirus and result in false positive measurements of antibody neutralization. Positive anti-SARS-CoV-2 nAb activity was defined as an anti-SARS-CoV-2 nAb titer >3 times greater than the titer of the same serum tested with the specificity control.

### Serum Samples

#### mRNA-1273 vaccine recipients

Neutralization activity was assayed in a total of 90 serum samples collected in a phase 1 trial^37,38^ (NCT04283461) that used the same mRNA-1273 vaccine, dose, and schedule as that used in the Moderna phase 3 trial^3^ (NCT04470427). Sera were collected from 30 vaccine recipients at day 0, day 29 (4 weeks post-first dose), and day 57 (4 weeks post-second dose). Samples were stored at −80°C, thawed, and heat-inactivated for 30 minutes (Duke) or 60 minutes (Monogram) at 56°C. Heat-inactivated samples were stored at 4°C (Duke) or at −80°C (Monogram) until assayed.

#### Convalescent patients

Neutralization activity was also assayed in a total of 248 serum samples collected in HVTN 405/HPTN 1901 (NCT04403880), an observational cohort study that enrolled individuals 18 years or older who had a positive SARS-CoV-2 test and recovered from a spectrum of infection and COVID-19 severity, from asymptomatic to requiring advance medical care in the intensive care unit, within 1-8 weeks of enrollment (if symptomatic). The laboratory was blinded to the clinical status of the donors but not to HIV-1 infection status. Only convalescent serum samples from HIV-1-uninfected individuals were used in the present analyses because antiretroviral drugs, which individuals with HIV may be taking, can inhibit replication of the HIV backbone of the pseudovirus used in the assay. Samples were stored at −80°C, thawed, and heat-inactivated for 30 minutes (Duke) or 60 minutes (Monogram) at 56°C. Heat-inactivated samples were stored at 4°C (Duke) or at −80°C (Monogram) until assayed.

#### WHO anti SARS-CoV-2 immunoglobulin International Standard (IS) sample

In December 2020, the WHO released a well-characterized international standard for the purpose of improving comparability of results among different assays in different laboratories and reducing interlaboratory variability of anti-SARS-CoV-2 antibody assays.^33^ As described in the user instructions provided by the National Institute for Biological Standards and Controls (NIBSC): “The First WHO International Standard for anti-SARS-CoV-2 immunoglobulin is the freeze-dried equivalent of 0.25 mL of pooled plasma obtained from eleven individuals recovered from SARS-CoV-2 infection. The preparation was evaluated in a WHO International Collaborative study. The intended use of the International Standard is for the calibration and harmonization of serological assays detecting anti-SARS-CoV-2 neutralizing antibodies. The preparation can also be used as an internal reference reagent for the harmonization of binding antibody assays. The preparation has been solvent-detergent treated to minimize the risk of the presence of enveloped viruses”.^39^

### Calibration approaches and evaluation of concordance

We discuss and compare three different approaches for the calibration of vaccine-induced neutralizing antibody responses measured by the Duke lab and the Monogram lab. The Monogram lab is used as the reference lab in Approaches 2 and 3 because vaccine-induced neutralizing antibody responses from 4 out of 5 of the USG-sponsored COVID-19 phase 3 vaccine trials are being assayed by, or are planned to be assayed by, the Monogram lab for correlates of protection assessments.

In Approach 1, the Duke and Monogram responses are calibrated to a common scale based on the WHO IS sample. For measurements from both labs the calibrated responses cID50 and cID80 in international units (IUs) per microliter (ml) are calculated as the observed ID50 or ID80 titers multiplied by the calibration factor, which is calculated as 1,000 (IU/ml) divided by the geometric mean, median or arithmetic mean ID50 or ID80 titers of the WHO IS sample that each lab measured in multiple vials. During the validation step, the calibration factor based on one of the above summary statistics that renders the highest agreement between calibrated responses from the two labs for the same set of samples is selected.

In Approach 2, both the Duke and Monogram responses are calibrated to a common scale using the Monogram lab as the reference based on methods described in Huang et al.^40^ Briefly, both readouts are assumed to share the same underlying true values but carry lab-specific measurement errors, following a bivariate normal distribution. Based on the assay validation experiments from both labs, a 30% coefficient of variation is assumed for the measurement error for ID50 and ID80 titers in both assays. The calibrated values are then calculated as the expected conditional means derived from the joint bivariate distribution as 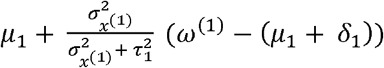 for the Monogram assay, where *μ*_1_ and 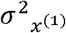 are estimated by the sample mean and variance of the log-transformed ID50 and ID80 titers of the convalescent samples measured by the Monogram assay, 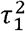 is the estimated measure error of the assay (= 0.3^2 = 0.09), *δ*_1_ is zero because the Monogram assay is considered as the reference, and *ω*^(1)^ is the log-transformed pre-calibrated titer of the vaccine sample measured by the Monogram assay. Similarly, the calibrated values for the Duke assay are calculated as 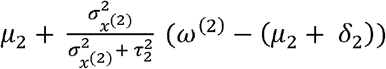, where all parameters are estimated using data from the Duke assay as is done for the Monogram assay, except that *δ*_2_ is estimated by the average of the log-transformed difference between the Duke and Monogram assay readouts of the convalescent samples.

In Approach 3, the Duke responses are calibrated using the relationship between Duke and Monogram readouts of the convalescent samples. Specifically, a linear regression model is used to regress the Monogram readouts on the Duke readouts; the calibrated Monogram scale value of a Duke sample is calculated by plugging in the observed Duke vaccine readout as the predictor in the regression model.

The concordance correlation coefficient (CCC)^41^ is used to quantify the level of agreement between the calibrated values of the two labs using samples measured by both labs. The overall CCC is then calculated as the average time-specific CCC using an inverse hyperbolic tangent transformation for stability. For both Approaches 2 and 3, to account for the statistical variability in the calibrated responses, 1000 bootstrap samples of the convalescent paired sample dataset are drawn with replacement to estimate the precision of the estimated CCC.

## Data Availability

Access to patient-level data and supporting clinical documents by qualified external researchers is available upon request and subject to review.

## Acknowledgments

We thank the HVTN 405/HPTN 1901 trial participants, the Moderna Phase 1 trial participants, the protocol teams, site staff, and investigators. We thank Hua Zheng for early analyses of the convalescent sample data and assay validation data. We also thank the USG Immune Assay Capacity team and leadership, Christopher Houchens, Ruben O. Donis, Lakshmi Jayashankar, Karen Martins, Flora Castellino, Brett Chromy, Mark Delvecchio, Tom Hu, Corey Hoffman, Evan Sturtevant, Robert A. Johnson, Xiaomi Tong, Tremel Faison, and Leah Watson from BARDA, Richard Koup and Janet Lathey from NIH, Chris Roberts from DMID at NIAID, Najaf Shah and Andrew Li from Boston Consulting Group, and Kaia Lyons and Kelsey Engel from Duke University. This research has been funded by the National Institute of Allergy and Infectious Diseases, National Institutes of Health (UM1AI068635, UM1AI068614, and UM1AI068618). This research has also been funded in whole / part by a federal contract from the National Institute of Allergy and Infectious Diseases, National Institutes of Health: Collaborative Influenza Vaccine Innovation Centers (CIVICs) Component A: Vaccine Center (Contract Number: 75N93019C00050).

The mRNA-1273 phase 1 study was sponsored and primarily funded by the National Institute of Allergy and Infectious Diseases (NIAID), National Institutes of Health (NIH), Bethesda, MD; in part with federal funds from the NIAID under grant awards UM1AI148373, to Kaiser Washington; 5 UM1AI148576, UM1AI148684, and NIH P51 OD011132, to Emory University; NIH AID AI149644, and contract award HHSN272201500002C, to Emmes. The study was conducted in collaboration with ModernaTX and funding for the manufacture of mRNA-1273 phase 1 material was provided by the Coalition for Epidemic Preparedness Innovation.

## Author Contributions

Conceptualization: Y.H., O.B., P.B.G., C.J.P., and D.C.M. Formal analysis: J.J.K. and Y.H. Investigation: T.W., S.C., M. S.-K., C.M., A.E., R.P., J.H., K.G., and S.K. Supervision: Y.H., L.C., P.B.G., C.J.P., and D.C.M. Funding acquisition: Y.H., P.B.G., M.J.M., L.C. Writing – Original draft: Y.H., O.B., and L.N.C. Writing – Review and editing: all authors.

## Ethics Declaration

The work described here was approved by the Duke University Health System Institutional Review Board (Duke University) through protocol IDs Pro00093087 and Pro00105358.

