## Supplementary Material for "Calibration of Two Validated SARS-CoV-2 Pseudovirus Neutralization Assays for COVID-19 Vaccine Evaluation"

**Figure S1: Correlations of Observed (A, B) ID50 and (C, D) ID80 titers between Day 29 and Day 57 vaccinee samples measured by (A, C) Duke and (B, D) Monogram.**

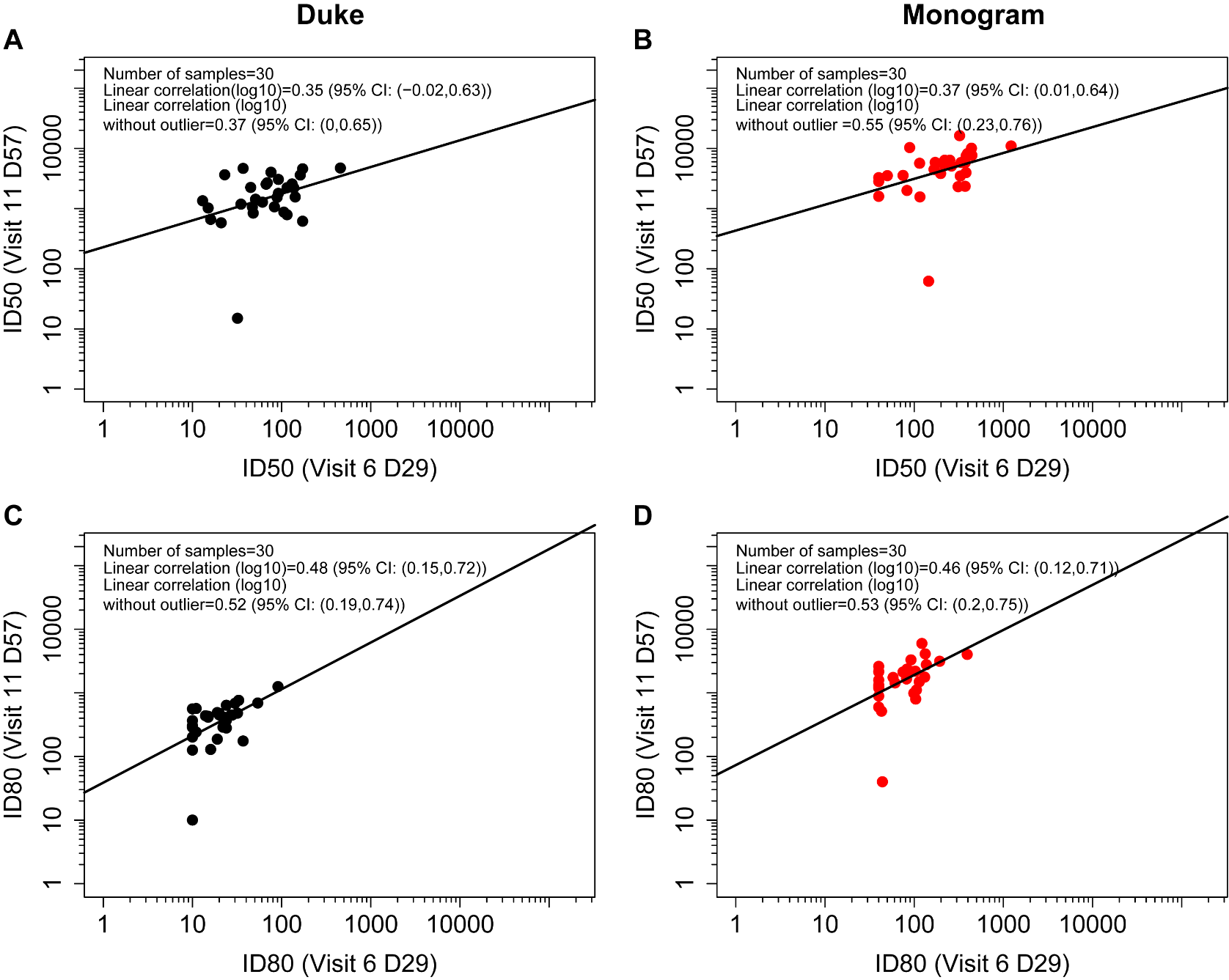

**Figure S2: Comparisons of the performance of Approach 1 based on calibration factor calculated using the (A, D) arithmetic mean, (B, E) geometric mean, or (C, F) median and (A-C) ID50 or (D-F) ID80 titers.** Axis labels cID50, cID80: ID50, ID80 titers calibrated to the WHO International Standard, expressed in International Units per ml (IU/ml).

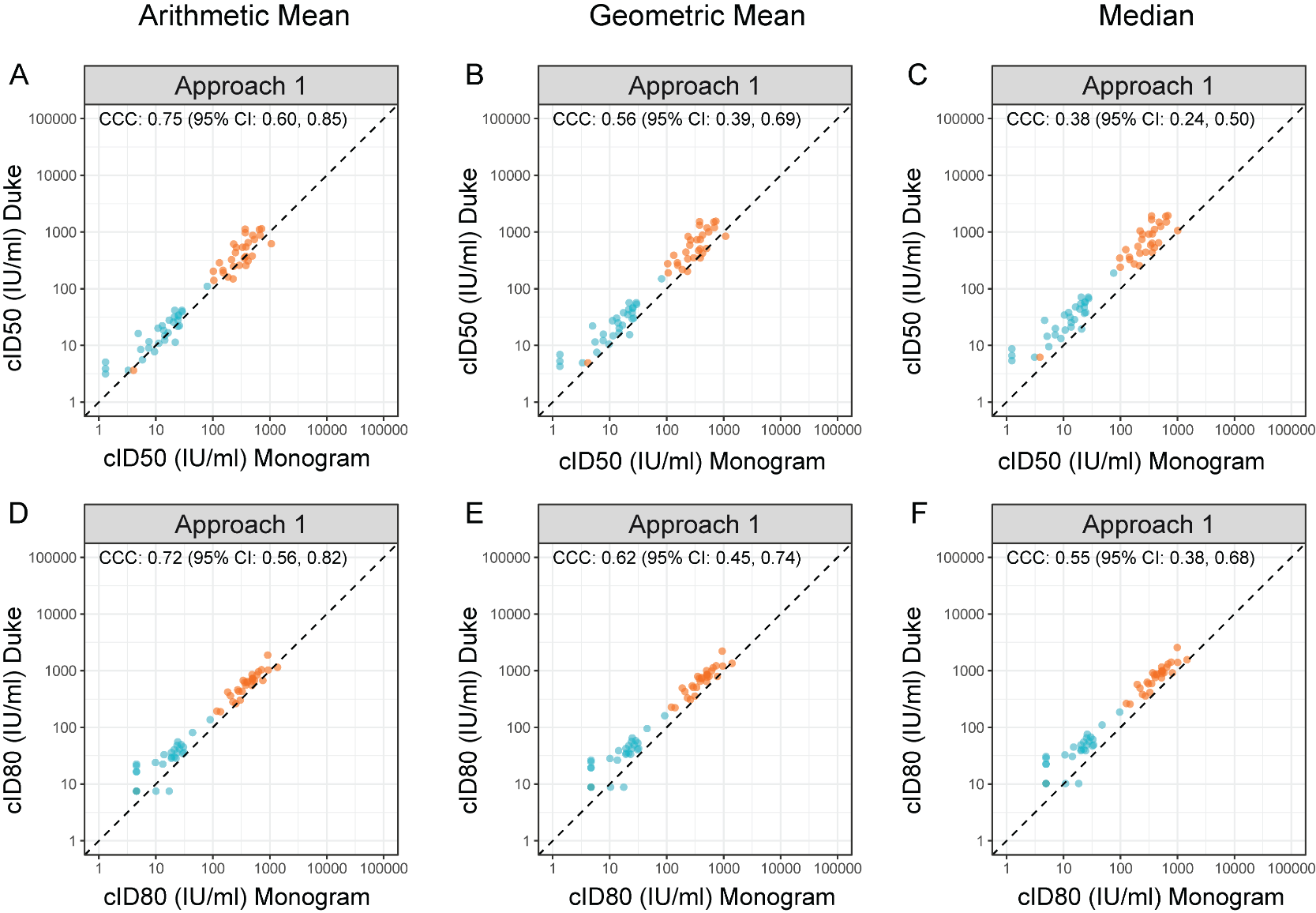

**Figure S3: Scatterplots of observed (A) ID50 and (B) ID80 Duke vs. Monogram of the 248 convalescent patient samples.** The solid line is the best fit linear regression line. The dashed line is the x=y diagonal line.

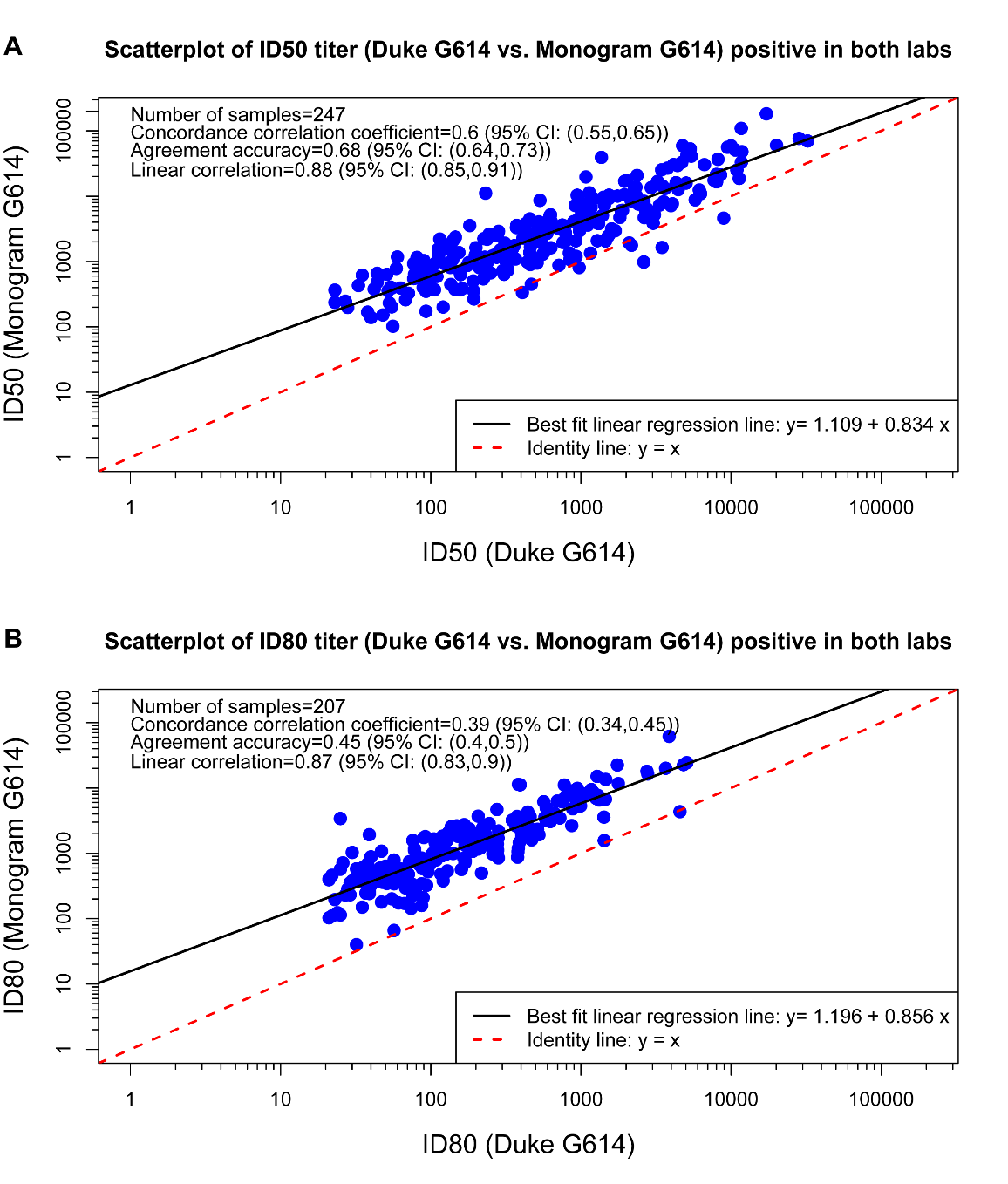

**Table S1. Approach 1 Calibration Factors and Conversation Factors.** The calibration factors are calculated as 1,000 (IU/mL) divided by the arithmetic mean, median or geometric mean ID50 or ID80 titers of the WHO IS sample that each lab measured on multiple vials. The conversion factors are the ratio of the calibration factors between the two labs.

| **Calibration** | **Lab** | **Titer** | **Lab-specific calibration factor** | **Between-lab (Duke vs. Monogram) conversion factor** |
| --- | --- | --- | --- | --- |
| Arithmetic mean | Duke | ID50 | 0.2418380 | 3.7 |
|  | Monogram |  | 0.0652635 |  |
|  | Duke | ID80 | 1.5010507 | 6.6 |
|  | Monogram |  | 0.2281074 |  |
| Geometric mean | Duke | ID50 | 0.3282240 | 4.9 |
|  | Monogram |  | 0.0666778 |  |
|  | Duke | ID80 | 1.7646021 | 7.5 |
|  | Monogram |  | 0.2347583 |  |
| Median | Duke | ID50 | 0.4129672 | 6.7 |
|  | Monogram |  | 0.0619848 |  |
|  | Duke | ID80 | 2.0449898 | 8.3 |
|  | Monogram |  | 0.2461538 |  |

**Table S2. Advantages and Disadvantages of Calibration Approaches 1-3.**

|  | **Advantages** | **Disadvantages** |
| --- | --- | --- |
| **Approach 1** | - Intuitive - Does not require labs running a panel of independent (convalescent) samples | - Requires running a sufficient number of vials of the WHO IS sample - Does not incorporate individual-level correlations of assay readouts between labs, hence individual-level calibration may suffer from lack of accuracy. - Arbitrary choice* of methods for calculating the calibration factor. |
| **Approach 2** | - Best performance - Simple to implement when necessary paired data are available | - Requires running a panel of independent (convalescent) samples. - Relies on the bivariate normal assumption and additive measurement error for assay readouts from the two labs, which may not be true. |
| **Approach 3** | - Intuitive | - Requires running a large panel of independent (convalescent) samples. - Does not account for measurement error in the assays |
